# Pre-pandemic blood profiles predict COVID-19 hospitalization and death a decade later

**DOI:** 10.64898/2026.05.27.26354230

**Authors:** Laurence A. Jacobs

**Affiliations:** Center for Molecular Cardiology, University of Zurich, Switzerland; Center for Complexity Sciences, National University of Mexico, Mexico City, Mexico

**Keywords:** COVID-19, prodromal risk, UK Biobank, proteomics, IL-6, IL-1, inflammation, pandemic preparedness

## Abstract

COVID-19 risk scores developed during the pandemic relied on measurements contemporaneous with infection, leaving unresolved whether the metabolic and inflammatory vulnerability they capture pre-existed as a stable trait or was triggered by acute illness. Here, using 501,946 UK Biobank participants whose blood was drawn between 2006 and 2010—at least ten years before SARS-CoV-2 emerged—we show that baseline proteomic and metabolic profiles predict both COVID-19 hospitalization (2,783 events; C* = 0.676 [0.666–0.686]) and COVID-19 mortality (1,564 deaths; C* = 0.730 [0.701–0.760]) from parsimonious, regularized feature sets. The IL-1 pathway index (xIL1, +0.093) was independently selected for hospitalization but not mortality, while the IL-6 trans-signaling index (xIL6, +0.040) was selected for mortality but not hospitalization—a differential pathway weighting corroborated by independent Light-GBM/SHAP analysis and mirroring the subsequent success of tocilizumab (anti-IL-6R) and the limited efficacy of anakinra (anti-IL-1R) in reducing COVID-19 mortality in randomized trials conducted years later. The mortality model was additionally characterized by central adiposity (waist-hip ratio, +0.386), a respiratory compromise index (xRSP, +0.149), and prodromal cardiovascular disease (pCVD, +0.246). These findings establish that vulnerability to a novel pathogen is, in substantial part, a pre-existing and measurable prodromal state, with implications for pandemic preparedness and population-level risk stratification.

## Introduction

The observation that pre-existing cardiometabolic disease amplifies COVID-19 severity was among the earliest and most replicated findings of the pandemic [1–3]. Obesity, diabetes, chronic kidney disease, and hypertension were rapidly identified as risk factors for hospitalization and death [4–6], and IL-6 emerged as the dominant cytokine mediator of the life-threatening hyperinflammatory response [7, 8]. Risk scores developed during the pandemic—notably QCOVID [9] and the 4C Mortality Score [10]—used measurements obtained at or after infection. What remained unclear was whether these risk factors merely tagged prevalent disease or captured a deeper layer of physiological vulnerability—one that exists as a measurable trait years before any encounter with the pathogen.

This question has practical significance beyond COVID-19 [11]. If vulnerability to a novel respiratory pathogen is partly determined by a stable metabolic and inflammatory phenotype, then population-level proteomic profiling could identify high-risk individuals *before* the next pandemic arrives, enabling pre-emptive shielding, vaccine prioritization, and targeted metabolic intervention. If, conversely, risk is dominated by acute-phase responses to infection itself, baseline profiling adds little.

We exploit a natural experiment to distinguish these possibilities. The UK Biobank collected blood samples and detailed phenotyping from approximately 502,000 participants between 2006 and 2010 [12]. Olink proteomic profiling of stored plasma was completed in 2023 [13, 14], providing measurements of *∼*3,000 proteins at a time point that pre-dates the SARS-CoV-2 pandemic by at least a decade. COVID-19 hospitalization and death data were subsequently linked through Hospital Episode Statistics and death registries [15].

The temporal separation between baseline measurement (2006–2010) and outcome (2020–2023) eliminates reverse causation from acute infection, producing an effectively prospective cohort study of prodromal vulnerability to a pathogen that did not yet exist.

## Results

### Study population and endpoints

From the full UK Biobank cohort of 501,946 participants with complete baseline data (median age 58.0 years [IQR 50.0–63.0]; 54.4% female; 88.1% White British), we defined two endpoints: COVID-19 hospitalization (COV), identified from Hospital Episode Statistics using ICD-10 codes U07.1 and U07.2 (2,783 events, 0.55%); and COVID-19 mortality (dCOV), identified from death registrations with U07 recorded as the underlying cause of death (1,564 deaths, 0.31%). Baseline assessments took place between March 2006 and October 2010 (median January 2009). Follow-up extended from the date of baseline assessment to death, loss to follow-up, or 13 June 2024, whichever came first (median follow-up 14.6 years; 7.14 million person-years). A total of 28,839 participants died before 1 January 2020 and were therefore not at risk for COVID-19 outcomes.

The median interval between baseline blood draw and COVID-19 death was 12.2 years (IQR 11.3–13.0; range 9.8–16.5). COVID-19 deaths spanned from 5 March 2020 to 13 June 2024, with a median date of 14 January 2021, placing the majority of events in the pre-vaccine and early-vaccine era.

Participants who died from COVID-19 were older at baseline (median age 62 vs. 58 years), more likely to be male (56.4% vs. 45.6%), had higher BMI (28.8 vs. 26.7 kg/m^2^), larger waist circumference (98.2 vs. 90.0 cm), and heavier smoking exposure (29.5 vs. 19.0 pack-years) than survivors.

### Prodromal prediction of COVID-19 hospitalization

The COV model was constructed using a four-stage pipeline: (1) 76 literature-informed prior features drawn from published COVID-19 risk factor studies entered the candidate set directly; (2) univariate discovery screening of the remaining features, retaining those with C* *>* 0.53 (none exceeded this threshold, indicating that the literature priors already captured the available signal); (3) LightGBM-based feature ranking with 5-fold cross-validation, selecting the top 25 features; and (4) *L*_2_-regularized logistic regression with balanced class weights and Platt recalibration on the selected feature set.^1^

The final COV model achieved C* = 0.676 (95% CI 0.666–0.686; Brier score 0.0055; calibration slope 0.97) with 25 selected features. The model was dominated by prodromal disease scores, respiratory physiology, and socioeconomic factors (Table 1): prodromal respiratory disease (pRD, +0.231) was the strongest predictor, followed by waist-hip ratio (+0.148), household income (−0.146), Townsend deprivation index (+0.122), FEV_1_ (+0.121), the respiratory compromise index (xRSP, +0.119), and prodromal cardiovascular disease (pCVD, +0.112).

**Table 1:**
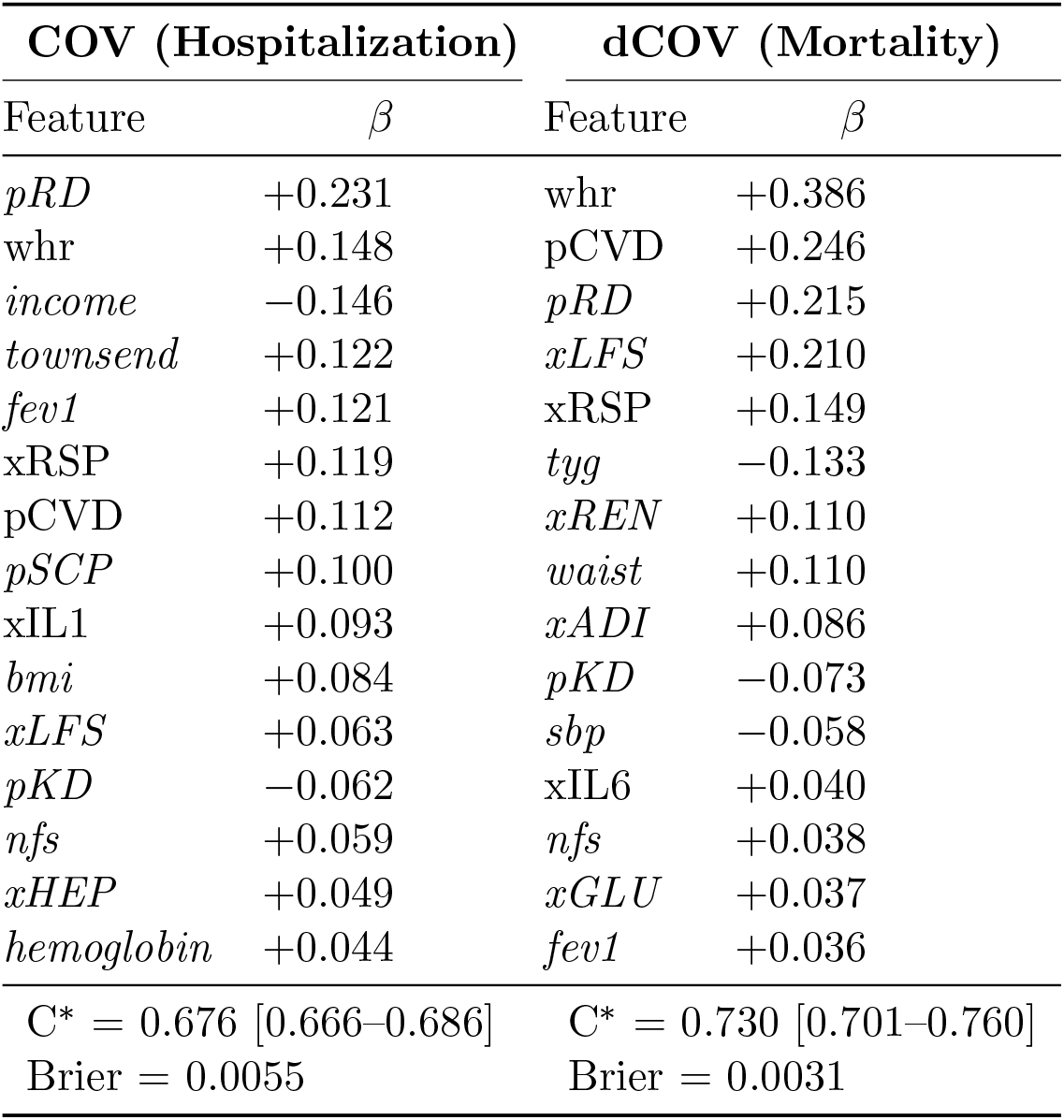
Selected features for COVID-19 hospitalization (COV) and mortality (dCOV). Coefficients are standardized log-odds ratios from *L*_2_-regularized logistic regression with balanced class weights and Platt recalibration. The dCOV model was refit after exclusion of two collinear liver fibrosis indices (FIB-4 and APRI; see Sensitivity analyses). Features are listed in order of absolute coefficient magnitude; top 15 shown for each endpoint. Cross-metric indices (x-prefix) are composite proteomic scores; prodromal scores (p-prefix) are regularized predictors of future disease incidence. Features unique to one model are italicized.

The IL-1 pathway index (xIL1, +0.093) was independently selected, while the IL-6 trans-signaling index was not retained in the final model. Prodromal sarcopenia (pSCP, +0.100) and the lifestyle index (xLFS, +0.063) contributed independently, indicating that vulnerability to COVID-19 hospitalization reflects convergent prodromal dysfunction across respiratory, cardiovascular, and inflammatory systems.

### Prodromal prediction of COVID-19 mortality

The dCOV model, constructed identically from 74 literature priors (no additional features exceeded the discovery threshold), achieved C* = 0.730 (95% CI 0.701–0.760; Brier score 0.0031; calibration slope 0.97) with 23 selected features after exclusion of two collinear liver fibrosis indices (see Sensitivity analyses).

The feature profile differed markedly from the hospitalization model. Central adiposity dominated (waist-hip ratio +0.386, waist +0.110) [16, 17], together with prodromal cardiovascular disease (pCVD, +0.246), prodromal respiratory disease (pRD, +0.215), and the lifestyle index (xLFS, +0.210). The respiratory compromise score (xRSP, +0.149), renal dysfunction index (xREN, +0.110), and adipokine index (xADI, +0.086) contributed independently, painting a picture of convergent multi-organ prodromal dysfunction.

The IL-6 trans-signaling index (xIL6, +0.040) was retained in the mortality model despite its modest coefficient, while the IL-1 pathway index (xIL1) was not selected in the top 25. This differential weighting—xIL1 selected for COV but not dCOV, xIL6 for dCOV but not COV in the *L*_2_-regularized model—is one of the central findings of this study and is corroborated independently by the LightGBM/SHAP analysis (see below).

### IL-1/IL-6 pathway asymmetry

A central finding is the differential weighting of inflammatory pathway indices across the two end-points (Table S1; Table 1). The IL-1 pathway index (xIL1, +0.093) was selected for hospitalization but not mortality; the IL-6 trans-signaling index (xIL6, +0.040) was selected for mortality but not hospitalization. In the *L*_2_-regularized model neither pathway enters the top 25 of the other end-point; in the LightGBM/SHAP analysis both pathways carry signal for both outcomes (Table S1), but with reversed dominance—xIL1 stronger for hospitalization (|SHAP| = 0.039 vs. 0.021), xIL6 stronger for mortality (|SHAP| = 0.059 vs. 0.032). The asymmetry is graded rather than absolute, but the rank ordering reverses cleanly between endpoints across both modeling frameworks.

This pathway-specific weighting—IL-1 more prominent for morbidity, IL-6 more prominent for mortality—aligns with the therapeutic evidence that emerged independently during the pandemic. Tocilizumab, an IL-6 receptor antagonist, reduced 28-day mortality in hospitalized COVID-19 patients in both the REMAP-CAP [18] and RECOVERY [19] trials, a finding confirmed by a WHO-sponsored meta-analysis of 27 trials [20]. Anakinra, an IL-1 receptor antagonist, showed more limited benefit [21], with efficacy restricted to a suPAR-defined hyperinflammatory subgroup rather than the broad hospitalized population. Canakinumab, a monoclonal antibody targeting IL-1*β*, did not reduce mortality in hospitalized patients [22].

That proteomic measurements made a decade before the pandemic recapitulate the pathway specificity of treatments tested years later—and do so through an unsupervised feature selection procedure with no COVID-19-specific tuning—provides strong evidence that xIL1 and xIL6 capture genuinely distinct and stable inflammatory biology, rather than artifacts of contemporaneous disease state.

The asymmetry is corroborated by the Stage 3 LightGBM models, which capture nonlinear effects and interactions that the final logistic regression cannot. In the GBM, xIL1 had mean |SHAP| = 0.039 for hospitalization but only 0.021 for mortality; xIL6 showed the reverse pattern (|SHAP| = 0.059 for mortality vs. 0.032 for hospitalization). That both a nonlinear ensemble and a linear model, applied to the same data with different inductive biases, agree on the rank ordering reinforces the conclusion that the asymmetry reflects biology rather than a statistical artifact of any single modeling framework.

### The hospitalization–mortality transition

Beyond the IL-1/IL-6 asymmetry, the two models reveal a broader shift in the biology of COVID-19 vulnerability as outcomes become more severe (Table 1).

#### From social to biological determinants

Socioeconomic factors (Townsend deprivation +0.122, household income −0.146) were prominent in the hospitalization model but absent from the top mortality predictors [23, 24]. The lifestyle index xLFS (+0.210, rank 4 for dCOV) likely absorbs part of the SES signal in the mortality model through its correlation with smoking, physical activity, and dietary patterns, but the explicit SES variables drop out: deprivation drives exposure and health-seeking behavior—who encounters the virus and who gets admitted—but once a patient is dying, the operative determinants are biological.

#### From respiratory-inflammatory reserve to central adiposity and cardiovascular substrate

The hospitalization model was led by prodromal respiratory disease (pRD, +0.231), with substantial contributions from FEV_1_ (+0.121), xRSP (+0.119), and xIL1 (+0.093)—a profile consistent with airway reserve and acute inflammatory tone determining who becomes ill enough to require admission. The mortality model shifts toward central adiposity (whr, +0.386; waist, +0.110), established cardiovascular substrate (pCVD, +0.246), and the lifestyle/metabolic/inflammatory composites (xLFS, +0.210; xREN, +0.110; xADI, +0.086; xIL6, +0.040)—a profile in which the underlying metabolic and cardiovascular ground state determines who, once severely ill, survives.

#### Respiratory physiology

The respiratory compromise index (xRSP) was selected for both hospitalization (+0.119) and mortality (+0.149), making it the only cross-metric index retained in both models. Prodromal respiratory disease (pRD) was the strongest single predictor of hospitalization (+0.231) and also a top-three predictor of mortality (+0.215). Subclinical respiratory compromise, measurable a decade before SARS-CoV-2 emerged, identifies individuals whose lungs cannot tolerate the additional insult of viral pneumonia.

#### Suppressor pairs and interpretive caution

The original 25-feature mortality model included two liver fibrosis indices, FIB-4 and APRI, which share AST and platelets as components (*r* = 0.957; PCA-1 explains 93.5% of joint variance with equal loadings). In the original model, FIB-4 appeared at +0.623 and APRI at −0.392—large coefficients of opposite sign that substantially deflated when FIB-4 was fit alone (+0.161). Dropping both reduced C* by only 0.002, confirming negligible independent contribution. The clean 23-feature model reported in Table 1 excludes both indices, eliminating the suppressor artifact while preserving discrimination. This analysis illustrates that coefficient magnitude in the presence of collinearity does not imply biological importance.

### Sensitivity analyses

Calibration was assessed by plotting observed event rates within deciles of predicted risk against mean predicted probabilities (Figure S2). Both models showed adequate calibration (calibration-in-the-large: COV *a* = −0.18, dCOV *a* = −0.16; calibration slope: COV 0.97, dCOV 0.97; integrated calibration index: COV 0.00075, dCOV 0.00058), with no systematic over- or underprediction across the risk spectrum. Decision curve analysis confirmed positive net benefit across the clinically relevant threshold range (COV: 0.5–5%; dCOV: 0.2–5%).

The FIB-4/APRI suppressor pair in the original dCOV model was investigated by ablation analysis on the full 501,946-participant cohort. The two indices are correlated at *r* = 0.957 (Spearman *ρ* = 0.73; PCA-1 explains 93.5% of joint variance with equal loadings). Removing APRI reduced FIB-4’s standardized coefficient from +0.623 to +0.161—a fourfold deflation characteristic of suppressor inflation—and dropping both indices reduced C* by only 0.002 (from 0.732 to 0.730). The clean 23-feature model excluding both indices is reported as the primary result.

To assess whether prediction required proteomic or prodromal score features, portable models were refit excluding all cross-metric (xMM) and prodromal (pFF) indices. The portable COV model (C* = 0.650) retained 8 features and the portable dCOV model (C* = 0.704) retained 8 features, both using only standard clinical measurements (Supplementary Table S2). The discrimination loss (ΔC* = 0.026 for COV, 0.026 for dCOV) indicates that the proteomic indices contribute biological insight—particularly the IL-1/IL-6 dissociation—rather than large increments of predictive accuracy beyond standard clinical measurements.

Additional planned sensitivity analyses for the peer-reviewed version include Cox proportional hazards confirmation with Schoenfeld residual tests, subgroup stratification by sex, age tertile, and ethnicity, and exclusion of the 28,839 participants who died before 1 January 2020.

## Discussion

The central finding of this study is that a parsimonious set of baseline blood measurements, collected a decade before SARS-CoV-2 emerged, predicts who will be hospitalized for and who will die from COVID-19, with discrimination (C* = 0.676 and 0.730, respectively) within the range of contemporaneous at-admission risk scores [9, 10], despite a fundamentally different prediction task: general-population baseline measurements taken a decade before infection, rather than clinical and laboratory measurements taken at or after hospital admission. The prediction is not merely a consequence of age and prevalent disease: the regularized models select proteomic composite indices (xIL1, xIL6, xRSP, xLFS) that carry independent information beyond demographics, comorbidities, and standard clinical biomarkers.

### Prodromal vulnerability as a pre-existing trait

The temporal design of this study—baseline measurement in 2006–2010, outcome in 2020–2023—establishes that vulnerability to a novel pathogen is, in substantial part, a stable trait measurable years in advance. This has direct implications for pandemic preparedness [11]. Population-scale proteomic or metabolic profiling could stratify populations into vulnerability tiers *before* the next pandemic, enabling pre-positioned shielding plans, tiered vaccine prioritization [25], and targeted metabolic interventions (weight management, glycemic control, smoking cessation, statin therapy) that reduce the substrate on which a novel pathogen acts.

### Pathway specificity as causal triangulation

The IL-6/IL-1 asymmetry provides a form of causal triangulation. The proteomic indices were constructed from baseline biology with no ref-erence to COVID-19. The therapeutic trials (REMAP-CAP, RECOVERY, SAVE-MORE) were conducted with no reference to prodromal proteomics. That both converge on the same pathway hierarchy—IL-6 more dominant for mortality, IL-1 more prominent for morbidity—strengthens the inference that the indices capture real inflammatory biology rather than statistical artifacts of high-dimensional feature selection. The convergence also suggests that IL-6 trans-signaling activation, even at subclinical baseline levels, identifies individuals who will mount a disproportionate cytokine response to a future inflammatory trigger [8].

### Central adiposity, not BMI

The dominance of waist-hip ratio (+0.386) with central adiposity indices in the mortality model reinforces the growing evidence that visceral fat distribution, not total body mass, is the operative COVID-19 risk factor [1, 5]. Visceral adiposity—with its attendant inflammatory, prothrombotic, and immunomodulatory consequences [16, 26]—is better captured by waist-based anthropometrics than by BMI [27]. This distinction, invisible to BMI-based risk stratification, was already present in the pre-pandemic baseline measurements.

### Limitations

The UK Biobank cohort is healthier, wealthier, and less ethnically diverse than the UK population [28], and the results may not generalize to populations with different comorbidity burdens or access to care. The Olink proteomic panel, while broad, does not capture all relevant immune mediators; a targeted cytokine panel measured closer to the pandemic might yield higher discrimination. The binary hospitalization endpoint does not distinguish severity of illness, and the mortality endpoint reflects both disease severity and treatment quality, which varied across pandemic waves. Finally, the models were developed and validated within a single cohort (internal cross-validation); replication in an independent biobank with pre-pandemic proteomics (e.g., the HUNT study or the Estonian Biobank) would strengthen the findings.

## Conclusion

Vulnerability to COVID-19—a disease caused by a pathogen that did not exist when the blood was drawn—is predictable from pre-pandemic proteomic and metabolic profiles with clinically meaningful discrimination. The pathway specificity of the prediction mirrors the therapeutic evidence that emerged independently during the pandemic. The differential biology of hospitalization versus mortality—from social to biological determinants, from respiratory and inflammatory reserve to central adiposity, cardiovascular substrate, and IL-6-mediated inflammation—argues for population-level molecular phenotyping as a component of pandemic preparedness infrastructure.

## Methods

### Study population

The UK Biobank recruited 502,460 participants aged 40–69 years between 2006 and 2010 across 22 assessment centers in England, Scotland, and Wales [12]. Baseline assessment included physical measurements, blood and urine sample collection, questionnaires, and linkage consent. The present analysis used 501,946 participants with complete baseline data.

### Outcome definitions

COVID-19 hospitalization (COV) was defined as at least one Hospital Episode Statistics (HES) record with ICD-10 diagnosis code U07.1 (COVID-19, virus identified) or U07.2 (COVID-19, virus not identified) in any diagnostic position (2,783 events, 0.55%). COVID-19 mortality (dCOV) was defined by the presence of ICD-10 code U07 as the underlying cause of death in death registration data linked to UK Biobank [15] (1,564 deaths, 0.31%). HES data were available through June 2024; death registrations through 13 June 2024. Follow-up was censored at death, loss to follow-up, or end of data linkage, whichever came first. Of the 1,564 COVID-19 deaths, the majority (median death date 14 January 2021) occurred during the pre-vaccine and early-vaccine period. A total of 28,839 participants who died before 1 January 2020 were not at risk for COVID-19 outcomes; these were retained in the analytic sample with censored follow-up times.

### Feature space

The candidate feature set comprised 248 variables spanning demographics, anthropometrics, vital signs, standard clinical biochemistry, hematology, lifestyle factors, socioeconomic indicators, prevalent disease flags, and medication use, plus two families of composite features derived from the PHRE platform.

#### Prodromal scores and cross-metric indices

Two families of composite features derived from the PHRE (Prodromal Health Risk Estimation) platform [29] were included as candidate predictors alongside standard clinical variables.

##### Prodromal scores (pFF)

Each prodromal score is a regularized predictor of a specific future disease, trained on the UK Biobank cohort using *L*_1_-penalized logistic regression. For a given endpoint (e.g., pDM for incident diabetes, pHT for incident hypertension, pKD for incident kidney disease), the score estimates the probability of developing that condition based on the participant’s current biomarker profile. Endpoint-specific feature exclusions prevent leakage—for example, HbA1c is excluded from pDM, and eGFR from pKD. Because prodromal scores for one disease may themselves be predictive of another (e.g., pDM predicts cardiovascular events), the full set of pFF scores is computed via damped self-consistent field (SCF) iteration (*α* = 0.5), analogous to Hartree–Fock mixing in computational physics, and iterated to convergence. The resulting scores capture multi-organ prodromal dysfunction in a compact, mutually adjusted representation.

##### Cross-metric indices (xMM)

Each cross-metric index is a composite score derived from Olink proximity extension assay (PEA) proteomic measurements [13, 14], constructed to capture a specific pathophysiological axis. For example, xIL1 aggregates proteins in the IL-1 signaling pathway, xIL6 captures IL-6/sIL-6R/gp130 trans-signaling, xRSP reflects respiratory compromise, and xNFL captures a dimension of systemic inflammation distinct from the interleukin axes. Cross-metric indices are standardized to zero mean and unit variance in the full cohort and are designed to be interpretable as pathway activation scores: a high xIL6 value indicates elevated baseline IL-6 trans-signaling, regardless of whether clinical symptoms are present. The composition of all cross-metric indices, including xIL1 and xIL6, was finalized in the PHRE v8.3 release prior to any COVID-19 outcome analysis; the panels were not refined in light of pandemic-era therapeutic trial results. Both pFF and xMM features are described in detail, including construction, validation, and the complete list of constituent proteins and endpoints, in Jacobs et al. [29].

### Model construction

Each endpoint was modeled using a four-stage pipeline:

#### Stage 1 (Literature priors)

For each endpoint, an *a priori* feature list was assembled from published risk factor studies. For COV, 76 features were included; for dCOV, 74 features.

#### Stage 2 (Univariate discovery)

The remaining features not in the literature prior list were screened univariately against the endpoint. Any feature exceeding C* *>* 0.53 was added to the candidate set. For both COV and dCOV, no additional features exceeded the discovery threshold.

#### Stage 3 (Feature selection)

LightGBM [30] gradient boosting with 5-fold cross-validation was used to rank features by importance; the top 25 were retained.

#### Stage 4 (Final model)

*L*_2_-regularized logistic regression was fitted on the 25 selected features with balanced class weights to address the low event rate (0.55% for COV, 0.31% for dCOV). Platt recalibration was applied to the predicted probabilities (COV: *a* = −5.17, *b* = 0.84; dCOV: *a* = −5.73, *b* = 0.76).

All features were standardized (zero mean, unit variance) before fitting, so coefficients represent the standardized log-odds-ratio contribution of each feature after mutual adjustment.

### Statistical analysis

Discrimination was assessed by C* (Harrell’s concordance statistic) with 95% bootstrap confidence intervals (1,000 replicates). Calibration was assessed by calibration-in-the-large (intercept) and calibration slope. Overall accuracy was assessed by the Brier score. SHAP (SHapley Additive exPlanations) [31] values were computed from the Stage 3 LightGBM models using TreeExplainer to visualize feature contributions at the individual level. Sensitivity analyses included Cox proportional hazards models, subgroup stratification by sex, age tertile, and ethnicity, and exclusion of pre-pandemic deaths.

## Data and code availability

UK Biobank data are available to approved researchers (https://www.ukbiobank.ac.uk). Analysis code is available from the corresponding author upon reasonable request.

## Ethics

UK Biobank has ethical approval from the NHS National Research Ethics Service (Ref: 11/NW/0382). All participants provided written informed consent.

## Acknowledgments

This research has been conducted using the UK Biobank Resource (https://www.ukbiobank.ac.uk) [12] under Application Number 596880. We thank all participants and the UK Biobank team for making this resource available. The Olink Explore 3072 proteomics data were generated by the UK Biobank Pharma Proteomics Project [13].

## Supplementary Material

**Figure S1:**
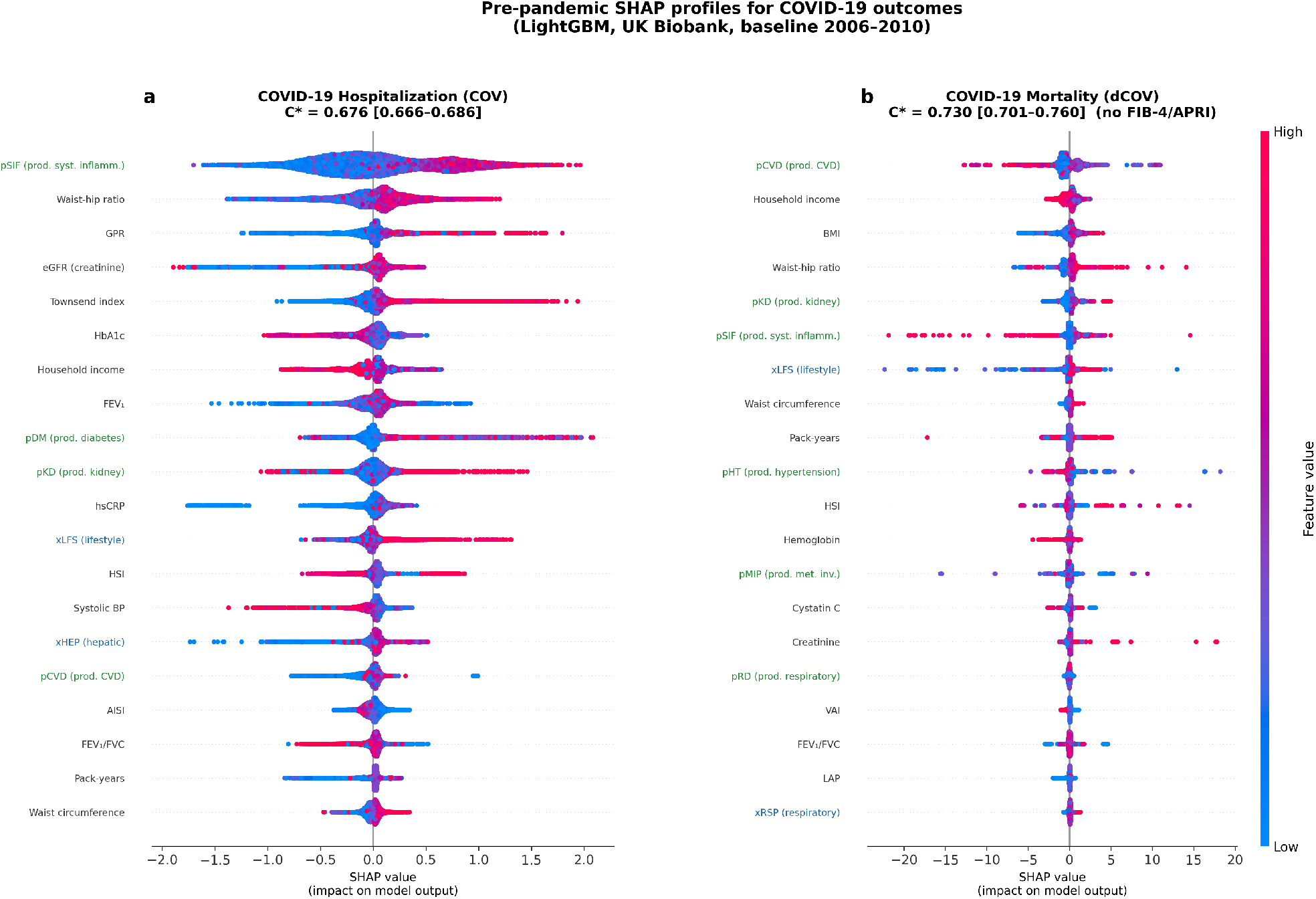
LightGBM SHAP beeswarm for COVID-19 hospitalization (COV, left) and mortality (dCOV, right). Each row corresponds to one feature and shows the per-individual SHAP contribution across the full 501,946-participant cohort. Points are coloured by feature value (red = high, blue = low). Features are ordered by mean |SHAP| (see Table S1). Note the reversed prominence of xIL1 (higher in COV) and xIL6 (higher in dCOV).

**Table S1:** LightGBM SHAP importance for COVID-19 hospitalization (COV) and mortality (dCOV). Mean absolute SHAP values from Stage 3 LightGBM models across the full 501,946-participant cohort. Features are ranked by mean |SHAP|; top 15 shown for each end-point. In the GBM, xIL1 shows higher importance for hospitalization (|SHAP| = 0.039) than mortality, while xIL6 shows higher importance for mortality (|SHAP| = 0.059) than hospitalization (|SHAP| = 0.032), corroborating the LR asymmetry.

| COV (Hospitalization) |  | dCOV (Mortality) |  |
| --- | --- | --- | --- |
| Feature | $\vert \text{SHAP}\vert $ | Feature | $\vert \text{SHAP}\vert $ |
| pCVD | 0.746 | pCVD | 1.449 |
| income | 0.173 | whr | 1.275 |
| townsend | 0.114 | pKD | 0.962 |
| xLFS | 0.095 | waist | 0.302 |
| pack_years | 0.092 | xADI | 0.192 |
| gpr | 0.077 | n_comorbidities | 0.170 |
| hscrp | 0.074 | xLFS | 0.154 |
| waist | 0.064 | tyg | 0.088 |
| lmr | 0.049 | fev1 | 0.084 |
| xRSP | 0.045 | xRSP | 0.079 |
| **xIL1** | **0.039** | bmi | 0.078 |
| xHEP | 0.039 | xGLU | 0.069 |
| pHRA | 0.033 | dbp | 0.061 |
| **xIL6** | **0.032** | **xIL6** | **0.059** |
| tyg_bmi | 0.031 | pSCP | 0.059 |
| GBM $C^* = 0.640$ | | GBM $C^* = 0.690$ | |

**Figure S2:**
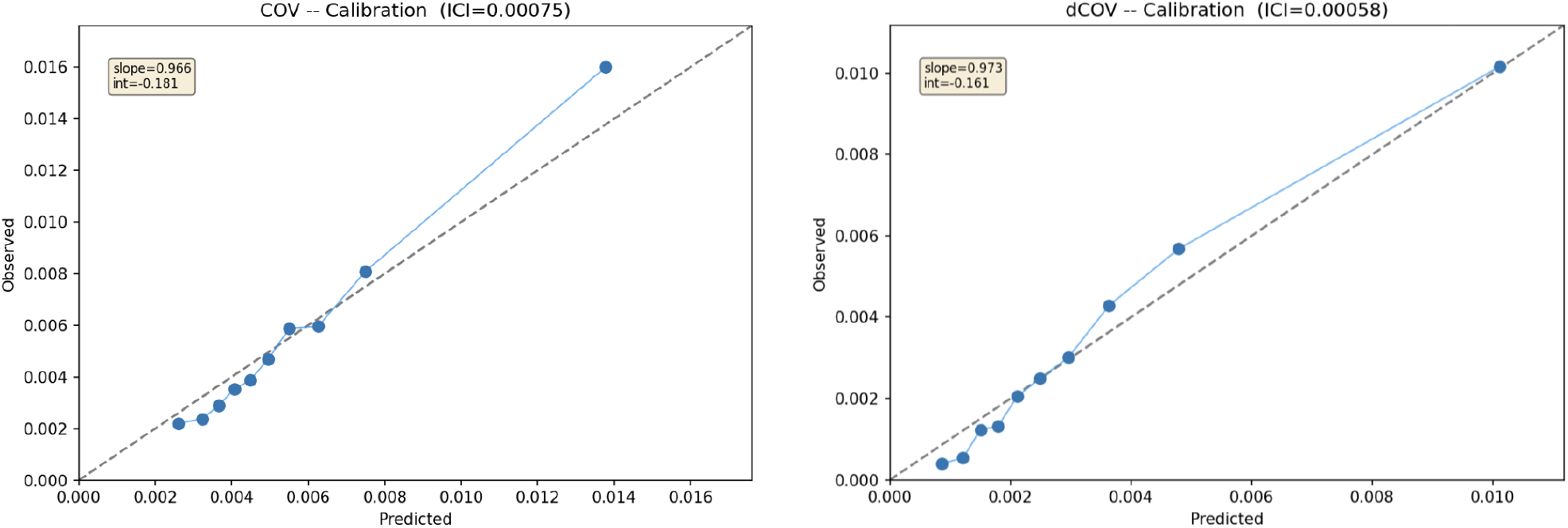
Calibration curves for COVID-19 hospitalization (COV: left) and mortality (dCOV: right). Observed vs. predicted event rates across deciles of predicted risk. The dashed line indicates perfect calibration. C* = 0.676; expected net benefit (ENB, a threshold-integrated measure of clinical utility [32]): COV = 7 × 10^−6^; dCOV = 4 × 10^−6^

**Table S2:** Portable model coefficients excluding proteomic and prodromal features. Models were refit using the same pipeline but excluding all cross-metric (xMM) and prodromal score (pFF) features, retaining only variables measurable with standard clinical assessments (COV: C* = 0.650; dCOV: C* = 0.704). Coefficients are standardized log-odds ratios from *L*_2_-regularized logistic regression with Platt recalibration. Predicted probability: 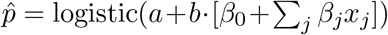, where *a* and *b* are Platt parameters (COV: *a* = −5.19, *b* = 0.91; dCOV: *a* = −5.75, *b* = 0.87) and *x*_*j*_ are standardized features.

| COV — Portable (8 features) |  | dCOV — Portable (8 features) |  |
| --- | --- | --- | --- |
| Feature | $\beta$ | Feature | $\beta$ |
| *intercept* | −0.144 | *intercept* | −0.270 |
| pack_years | +0.222 | waist | +0.791 |
| income | −0.185 | n_comorbidities | +0.247 |
| bmi | +0.180 | bmi | −0.237 |
| hba1c | +0.172 | fev1 | −0.179 |
| townsend | +0.146 | sbp | +0.136 |
| sbp | +0.054 | dbp | −0.122 |
| hemoglobin | +0.050 | hscrp | +0.038 |
| fev1 | −0.030 | hemoglobin | +0.004 |

## Footnotes

1 C* denotes Harrell’s concordance statistic, equivalent to the area under the receiver operating characteristic curve for binary outcomes.

## Notes

### Competing Interest Statement

The authors have declared no competing interest.

